# Rapid and Low-cost Sampling for Detection of Airborne SARS-CoV-2 in Dehumidifier Condensate

**DOI:** 10.1101/2020.10.08.20208785

**Authors:** Parikshit Moitra, Maha Alafeef, Ketan Dighe, Priyanka Ray, James Chang, Sai Sathish Ramamurthy, Xudong Ge, Dipanjan Pan, Govind Rao

## Abstract

Airborne spread of COVID-19 by infectious aerosol is all but certain. However, easily implemented approaches to assess the actual environmental threat are currently unavailable. We present a simple approach with the potential to rapidly provide information about the prevalence of SARS-CoV-2 in the atmosphere at any location. We used a portable dehumidifier as a readily available and affordable tool to collect airborne virus in the condensate. The dehumidifiers were deployed in selected locations of a hospital ward with patients reporting flu like symptoms which could possibly be due to COVID-19 over three separate periods of one week. Samples were analyzed frequently for both virus envelope protein and SARS-CoV-2 RNA. In several samples across separate deployments, condensate from dehumidifiers tested positive for the presence of SARS-CoV-2 antigens and confirmed using two independent assays. RNA was detected, but not attributable to SARS-CoV-2. Our results point to a facile pool testing method to sample air in any location in the world and assess the presence and concentration of the infectious agent in order to obtain quantitative risk assessment of exposure, designate zones as ‘hot spots’ and minimize the need for individual testing which may often be time consuming, expensive and laborious.

## Main

Since the emergence of the first case of coronavirus in Wuhan, China in December 2019, COVID-19 has infected over 36 million people and claimed 1,055,947 lives worldwide with a staggering number of 7,726,175 affected individuals in the US alone. The mortality rate is estimated to be around 1%, although these figures are not very accurate due to the lack of widespread testing and thereby under-reported. The unavailability of rapid testing has severely hampered efforts to manage the disease and assess its risk of transmission. Furthermore, uncertainty about its mode of spreading has created much perplexity and resulted in incoherent and constantly changing guidelines ^1^, creating public confusion and non-compliance. The case of mass infections from the Biogen conference, the Washington ChoiR^2^ and the Wuhan restaurant^3^ are concrete evidences of the ease with which social contact can spread the virus. Even as the World Health Organization (WHO)^4^ is evaluating the spread of SARS-CoV-2, understanding has evolved that the infection transmission mode is primarily respiratory through airborne transmission of aerosols^5^. Due to the shared similarities between SARS-CoV-2 and other coronaviruses like the Middle East Respiratory Syndrome (MERS-CoV) and Severe Acute Respiratory Syndrome (SARS-CoV), both of which were found to be airborne and could be potentially transmitted to long distances, it is essential to investigate this feature of the new virus and mitigate any plausible risks. Repeated aerodynamic analysis in hospitals^6-10^ and other indoor environments^11-14^ evidently present one commonality about COVID-19, the coronavirus does linger in the air and is highly infectious. Recent findings from a study conducted in a hospital ward further confirmed aerosol-based transmission of viable SARS-CoV-2 from air samples collected 2 to 4.8 m away from patients^15^. This highlights the requirement of an efficient yet facile technique to assess the presence of the virus in high density environments. Hence, understanding of our day-to-day exposure risk to these lethal bioaerosols is vital to implement near real-time interventions to prevent the spread of the virus as well as safeguard human health^16^. This is especially useful, as several reports indicating the spread of the virus through asymptotic and pre-symptomatic patients have surfaced^17,18^. A testing device thus placed in areas of high footfall and capable of bypassing individual testing is an effective way of controlling the spread of the deadly disease. Therefore, a simple, robust method, capable of providing rapid and accurate results on COVID-19 exposure would be extremely impactful to slow the spread of the disease and cater to community health at large.

We hypothesized that collecting condensate from the atmosphere could provide a simple means of assessing viral load in the surroundings. To this end, we set up four portable dehumidifiers at various test locations around a hospital ward at the University of Maryland Medical Center in Baltimore and obtained condensate samples for viral load analysis on different dates (**Fig. 1** and **Supplement Fig. 1**). The condensate was sampled at three separate time periods between June 29-July 5, July 22-August 10 and September 3-10, with the last set of samples being collected in viral transport medium (VTM). Our simple collection methodology aids in monitoring of SARS-CoV-2 virus in communal gatherings without the need for individual testing and could serve as a checkpoint for the dreaded second wave of coronavirus, even as the US saw a surge in the number of positive cases post relaxation of rules and policies governing social distancing over the Memorial Day weekend^19^. By employing this simple methodology (**Fig. 1**) to monitor the presence of SARS-CoV-2 especially in areas with high human footfall or mass gatherings, appropriate preventive measures can be adopted to identify & track possible hotspots and protect individuals from being infected while efforts to develop a vaccine against this deadly virus are ongoing.

**Figure 1.**
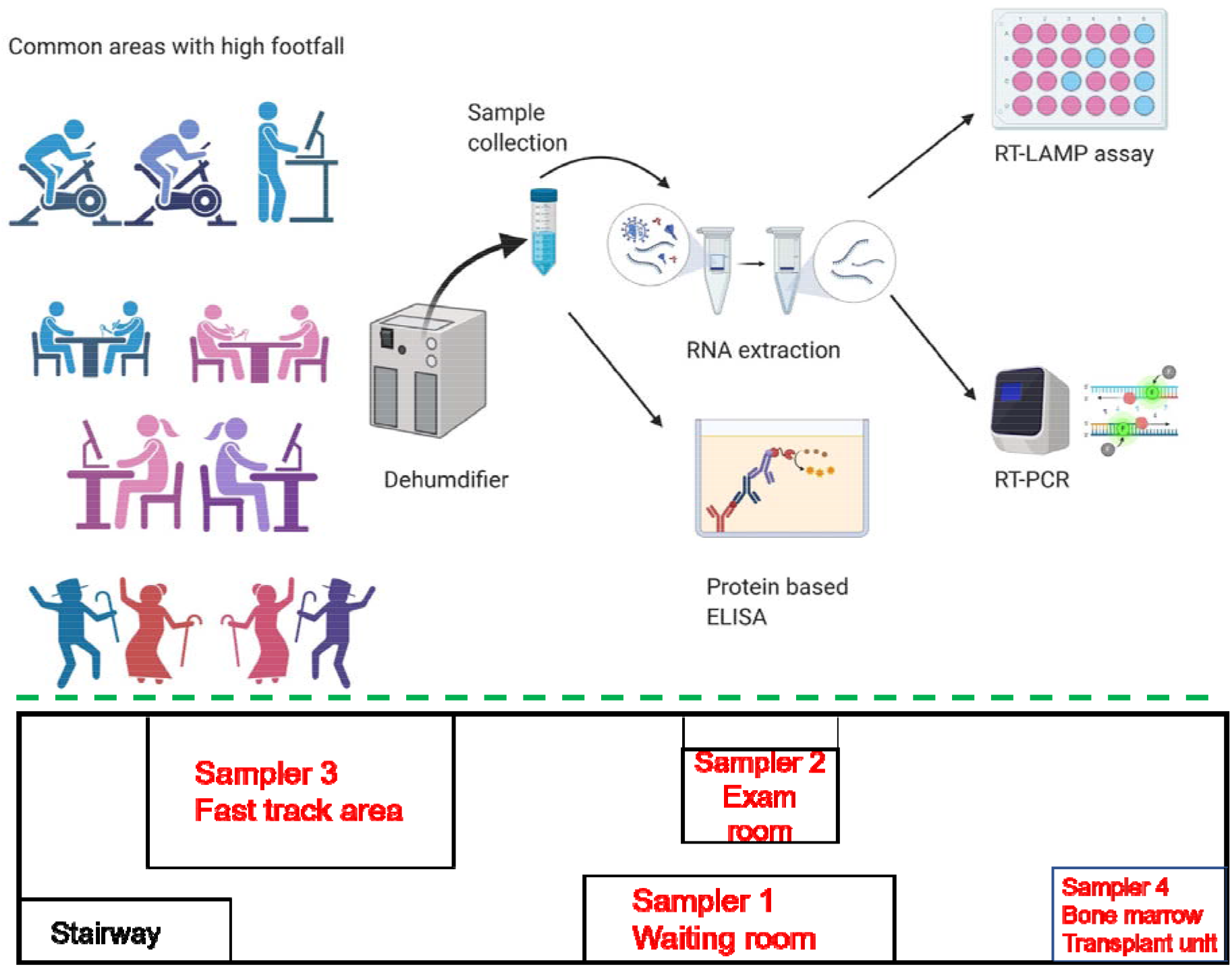
Schematic representation of sample collection and analysis for mass detection (top) and simplified layout of the hospital ward indicating the positions of various dehumidifiers during September 3-10, 2020 (bottom).

## Methods

Four identical 900 mL dehumidifiers (ICETEK B0863HNVNS from Amazon.com) were numbered 1 through 4 and deployed at the various sites indicated. These dehumidifiers use a muffin fan that draws room air past a peltier-cooled heat exchanger and deposits condensate into a tank underneath. The condensate tanks were sampled at 24 or 48 hour intervals and 50 mL samples were further processed for analysis. A three-prong approach was followed for the identification of viral load in the collected condensate samples. All samples were deactivated in a water bath set at 65 °C for 30 minutes, following which they were either stored at 4 °C for protein detection using a protein ELISA kit or aliquoted and freeze-dried for RNA based analysis employing commercially available RT-LAMP and RT-PCR kits. As a parallel detection technique, we employed a previously developed nanosensing platform from lanthanide doped carbon nanoparticles (LCNPs) which provide a distinct fluorescence response in presence of SARS-CoV-2.^20-22^ For samples collected in VTM, 50 mL of each sample was freeze dried and the residue redispersed in 2 mL of RNase free water and analyzed for the presence of RNA. Detailed procedure has been provided in the supporting information.

### Results and Discussion

Water samples collected between June 29 - July 5, 2020 were first analyzed through COVID-19 S-protein ELISA kit. All the experiments were carried out at room temperature and samples were tested in duplicates, the average value was then utilized to determine the final S-protein concentration. A calibration curve was initially generated using the known S-protein concentrations (**Fig. S2**) and S-protein in the samples was then estimated (**Table S1** and **Fig. 2**). Only one sample presented a detectable dose of the virus COVID-19 S-protein (dated July 5 from AED yellow zone by WGL214 door; S-protein concentration 2.61 ng/mL) while others were quite below the detection limit of the kit. Interestingly, the virus could be detected only when sampling was continued over the weekend in comparison with daily sampling protocol. This indicated the requirement of concentrate sampling for successful detection of viral spike protein. As a parallel detection technique, we employed a nanosensing platform that was previously developed in our laboratory.^20-22^ We tested 17 condensate (water) samples collected as described previously. The sensor consists of lanthanide doped carbon nanoparticles (LCNPs) provided a distinct fluorescence response towards the presence of SARS-CoV-2 specific viral protein (**Table S2**). Unsupervised Machine Learning algorithm (ML) was used to identify the presence of SARS CoV-2 using normalized fluorescence intensity.^20^ Inspired by the results, we decided to investigate the level of viral SARS-CoV-2 RNA in the water samples. RNA was extracted from all the samples (**Table S3**) and RT-LAMP and RT-PCR were performed to detect the presence of the viral SARS-CoV-2 RNA (**Fig. 3**).

**Figure 2.**
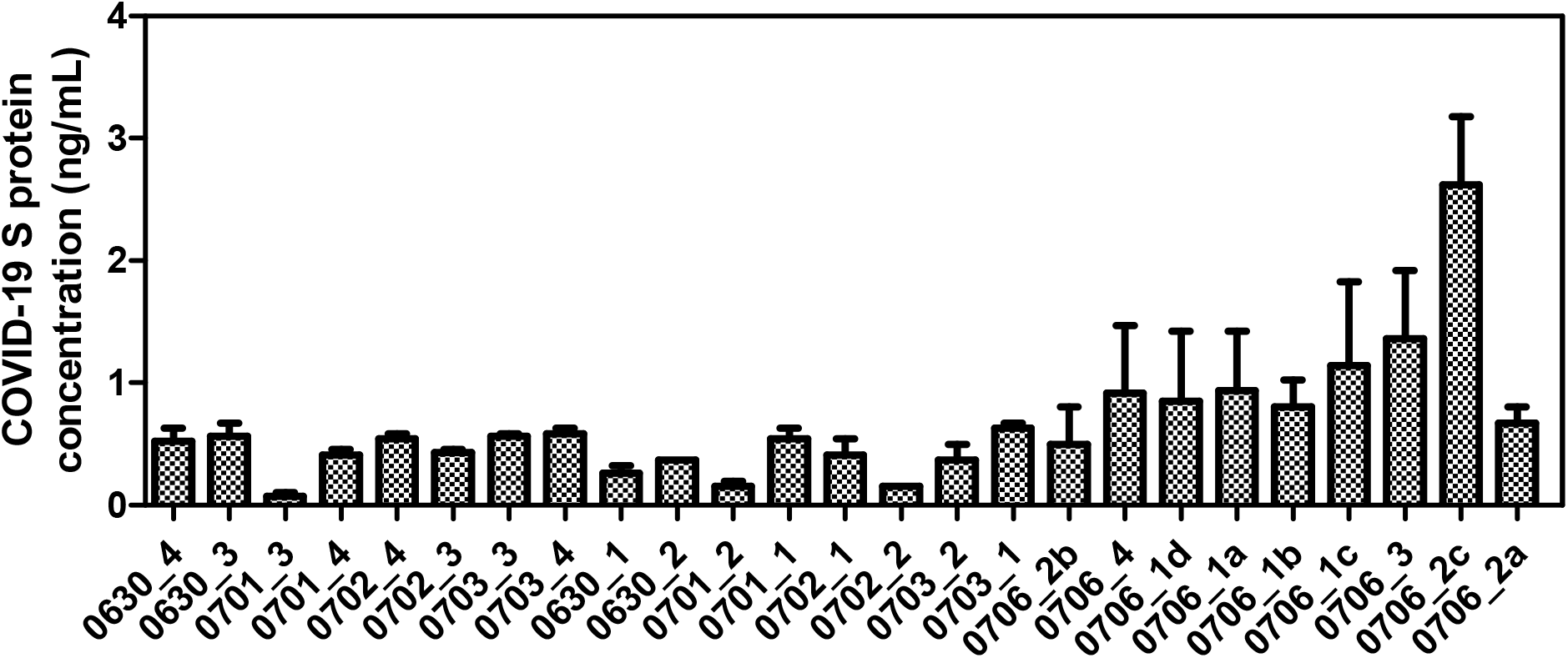
Concentration of COVID-19 S-protein as determined by the ELISA assay for the samples collected over the period of June 29 - July 5, 2020. The sample code starts with the date of sample collection from hospital followed by the dehumidifier number, i.e. 0630_4 indicates the water sample has been collected from dehumidifier number 4 on June 30, 2020.

**Figure 3.**
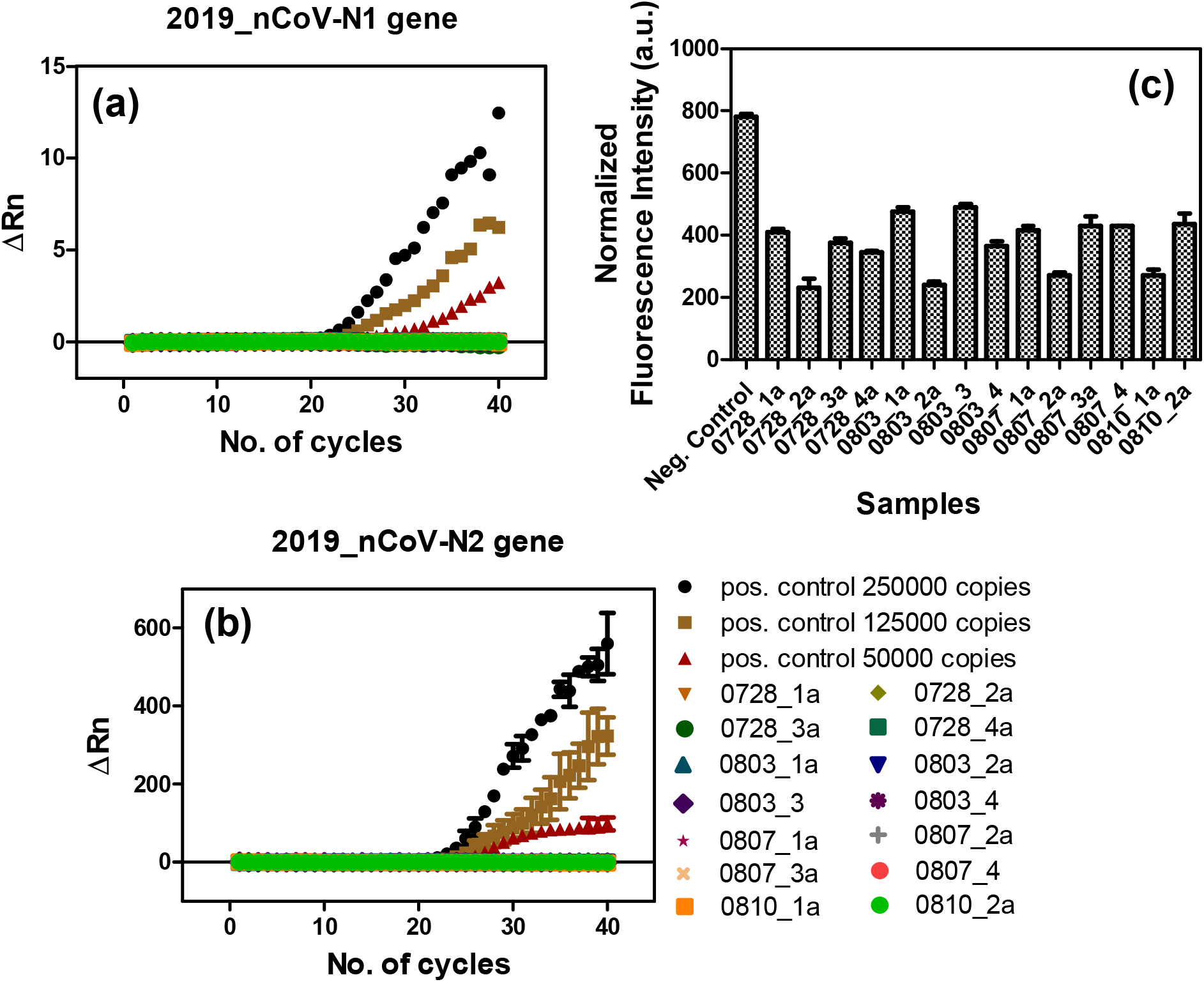
Determination for the presence of SARS-CoV-2 viral RNA by (a, b) RT-PCR and (c) RT-LAMP assay for the samples collected over the period of July 22 – August 10, 2020. The sample code starts with the date of sample collection from hospital followed by the dehumidifier number, i.e. 0728_1a indicates the water sample has been collected from dehumidifier number 1 on July 28, 2020.

Based on the results obtained using RT-LAMP and RT-PCR, no viral RNA was detected which we attribute to either the low detection limit of the methods used or due to deactivation or destabilization of SARS CoV-2 RNA in the dehumidifier chamber. In order to negate the possibility of viral destabilization in the sampling method, we added 50 mL VTM to the dehumidifier chamber. This was done to ensure the stability of the viral RNA in the condensate.

Following sample collection in VTM, although we were able to detect RNA in most of the samples (**Table S4**), RT-PCR (**Fig. 4a-b**) and RT-LAMP (**Fig. 4c**) showed absence of viral RNA. This negated our second inference regarding the destabilization of viral RNA in the sampler and alluded to the sensitive detection limits of the analysis which was rather difficult for samples with a possibly low viral RNA load as in the case of those collected from dehumidifiers. Interestingly, we were still able to detect the presence of S-protein close to the minimum detectable dose (**Fig. 4d**).

**Figure 4.**
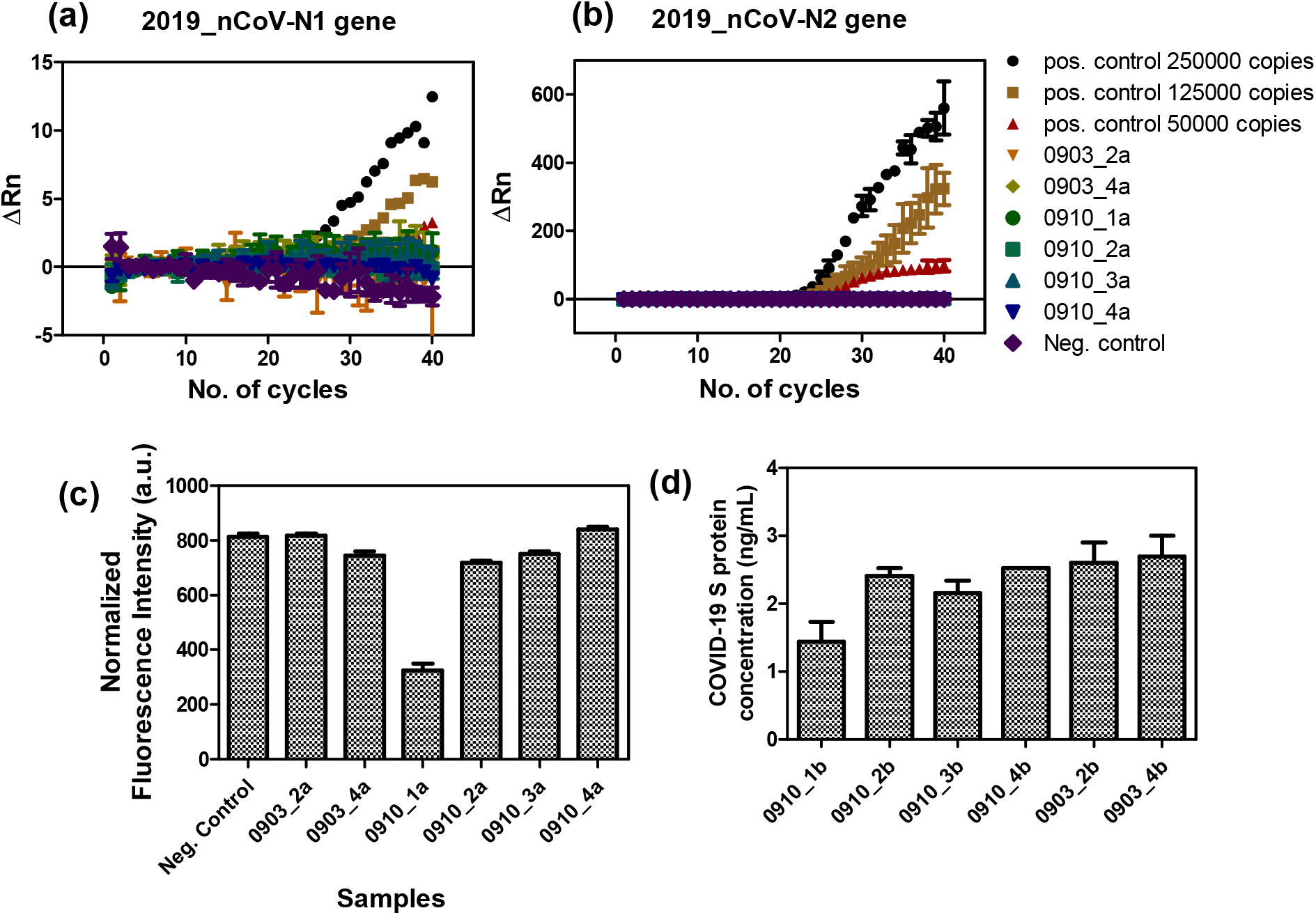
Determination for the presence of SARS-CoV-2 viral RNA by (a, b) RT-PCR and (c) RT-LAMP assay for the samples collected over the period of July 22 – August 10, 2020. The sample code starts with the date of sample collection from hospital followed by the dehumidifier number, i.e. 0903_2a indicates the water sample has been collected from dehumidifier number 2 on September 3, 2020.

The overall results of the study are summarized in **Table 1**. Most strikingly, SARS-CoV-2 viral protein was detected over some period in all the samplers. This could have implications for the efficacy of air filtration systems currently employed. Although airborne SARS-CoV-2 is widely implicated in the spread of COVID-19, there is great uncertainty over the precise mechanisms of exposure and susceptibility. The viral load in the atmosphere presumably fluctuates depending on the actual shedding by the infected person and their number. Our results cast a new light on this subject. We show that a simple technique of sampling condensate from a dehumidifier can provide evidence of airborne virus. Given the widespread use of air-conditioning equipment in homes and businesses worldwide, sampling their condensate provides a simple means of pool testing for virus presence analogous to those proposed for sewage monitoring. This approach also solves the major problem faced by conventional swab or saliva testing, where results can take several days. Antibody and point-of-care (POC) tests are more rapid but are geared towards individual patient testing and do not assess environmental airborne infection risk.

**Table 1.** Summary of results: Condensate samples collected during Phase I: June 29- July 5, July 22-August 10 and Phase II: September 3-10. Phase II samples included viral transport medium (VTM) in tank to stabilize any collected virus. RT-LAMP and RT-PCR analyses were performed on RNA isolated from samples; ELISA and Lanthanide array were performed directly on the samples.

| Number of Samples analyzed (Phase I without VTM) | Found positive using Protein ELISA | Found positive using Lanthanide array | Found positive using RT- LAMP | Found positive using RT-PCR |
| --- | --- | --- | --- | --- |
| 25 | 1<br>4% positive | 5<br>20% positive | Not detected | Not detected |

| Number of Samples analyzed (Phase II with VTM) | Found positive using Protein ELISA | Found positive using RT- LAMP | Found positive using RT-PCR |
| --- | --- | --- | --- |
| 8 | 5<br>62.5% positive | Not detected | Not detected |

Although RT-LAMP and RT-PCR based analyses did not detect the virus, this may be attributed to the dilution of the viral concentration in a large volume of media and inherent instability of the viral RNA in further processing steps used. Earlier reports on wastewater sampling and detection indicate the low concentration of the virus to be a major limitation^19^. RT-PCR has a Limit of Detection (LOD) of 6 copies/μL while RT-LAMP has a corresponding value of 0.75 copies/μL, both of which are way below the threshold of the milliliters range of detection required for sampling dilute solutions of viral load.

Since the Coronavirus is an enveloped virus, its recovery rate is substantially lower than that of non-enveloped viruses^23^. The major approaches to concentrate water samples include precipitation using polyethylene glycol (PEG), adsorption/elution, centrifugal ultrafiltration, aluminum hydroxide flocculation and electronegative filteration^24,25^. Recovery rates are also specific to the strain of the virus, their charge and hydrophobicity and partition to solids. Despite these study limitations, our primary results present a very novel method for air sampling in any resource limited settings across the globe. Coupled with sensitive and rapid assays that are being developed, there is the possibility of achieving near real-time sensing of SARS-CoV-2 in the atmosphere, thereby providing an actionable threat assessment.

## Conclusion

In the light of the recent pandemic, most countries are struggling to strike a balance between protecting their residents and keeping their economies from crashing further. In such unprecedented times, the world has witnessed overburdening of healthcare facilities and increased risk of transmission via healthcare workers and places with high human footfall. In an attempt to reduce the possibility of infection by adopting testing methods capable of producing effective and fast results in a cost effective manner, we have proposed herein a simple, facile and affordable testing method for areas with high population density or footfall by avoiding laborious and time consuming individual testing. The use of dehumidifiers in designated areas would allow for analysis of the collected condensate in a rapid and facile manner, thus allowing authorities to designate zones as ‘hot spots’ in case of a positive result. The method of sampling is both novel and effective, given the nature of transmission of coronaviruses and the unavailability of individual testing in many remote areas. With the much-dreaded possible approach of the second wave of the virus, this strategy would help in reducing the number of infected cases in a timely manner without exhausting public coffers and overwhelming the healthcare infrastructures especially in countries with scanty resources.

## Data Availability

All data are available upon request in addition to supplementary file.

## ACKNOWLEDGMENTS

We thank Dr. Bruce Jarrell for rapidly facilitating access to hospital sampling. The following reagent was deposited by the Centers for Disease Control and Prevention and obtained through BEI Resources, NIAID, NIH: Quantitative PCR (qPCR) Control RNA from Heat-Inactivated SARS-Related Coronavirus 2, Isolate USA-WA1/2020, NR 52347.

## AUTHOR CONTRIBUTIONS

Study concept and design by GR, JC, SSR and XG

Study design regarding the molecular testing of SARS-CoV-2 by DP, PM, KD and MA

Product analysis and method development (protein and nucleic acid) were done by DP, PM, MA, KD and PR

Sampling by JC, PM, KD and PR Sample preparations by KD and PR

Data analysis was done by PM, KD, PR, MA and DP Manuscript written by PM, PR, MA, KD, DP, SSR, GR

