## Supplement file for "Rapid and Low-cost Sampling for Detection of Airborne SARS-CoV-2 in Dehumidifier Condensate"

**Figures.**

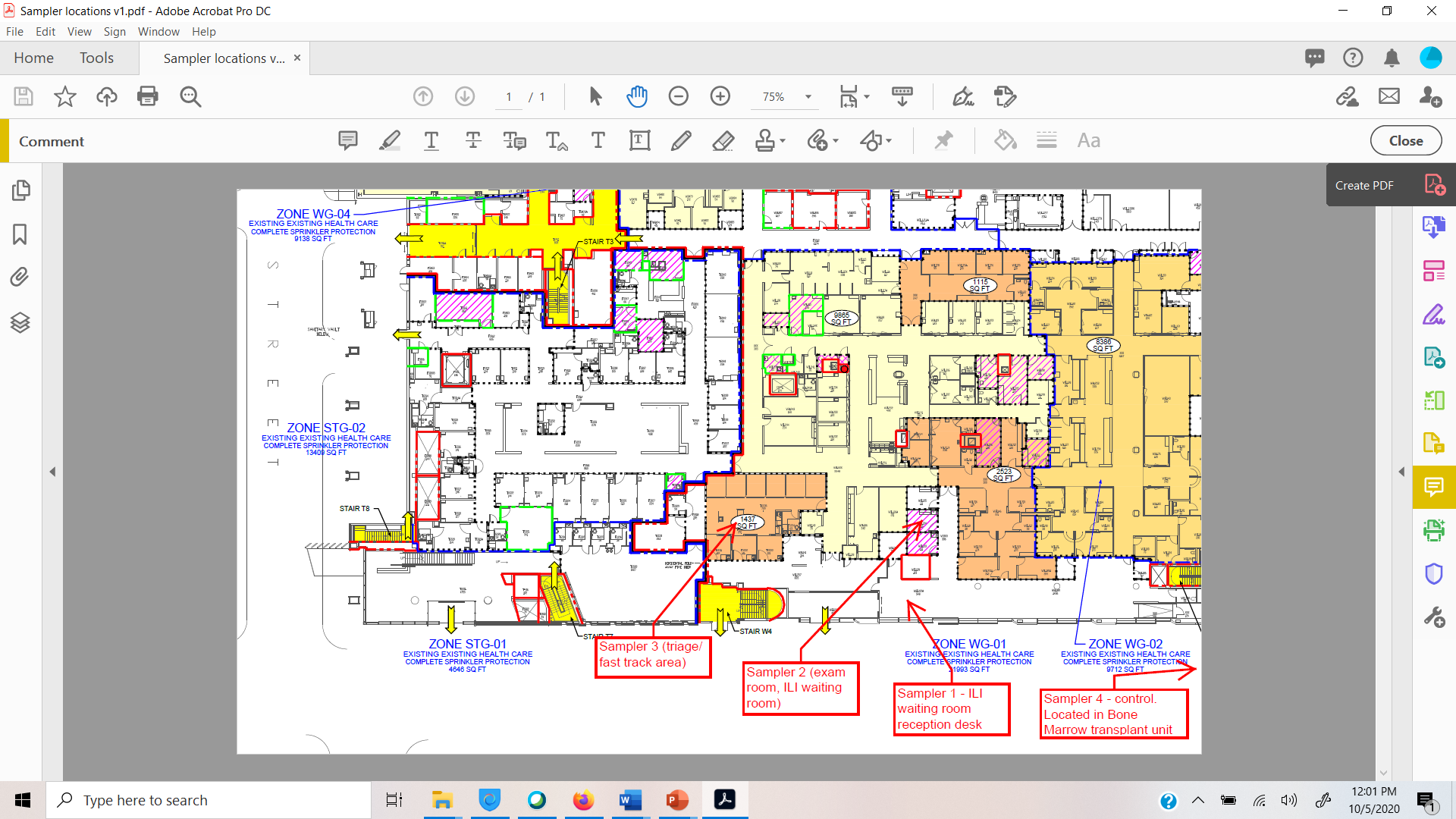

**Figure S1.** Detailed design map of the hospital wards showing the deployment of four humidifiers at different regions during September 3-10, 2020.

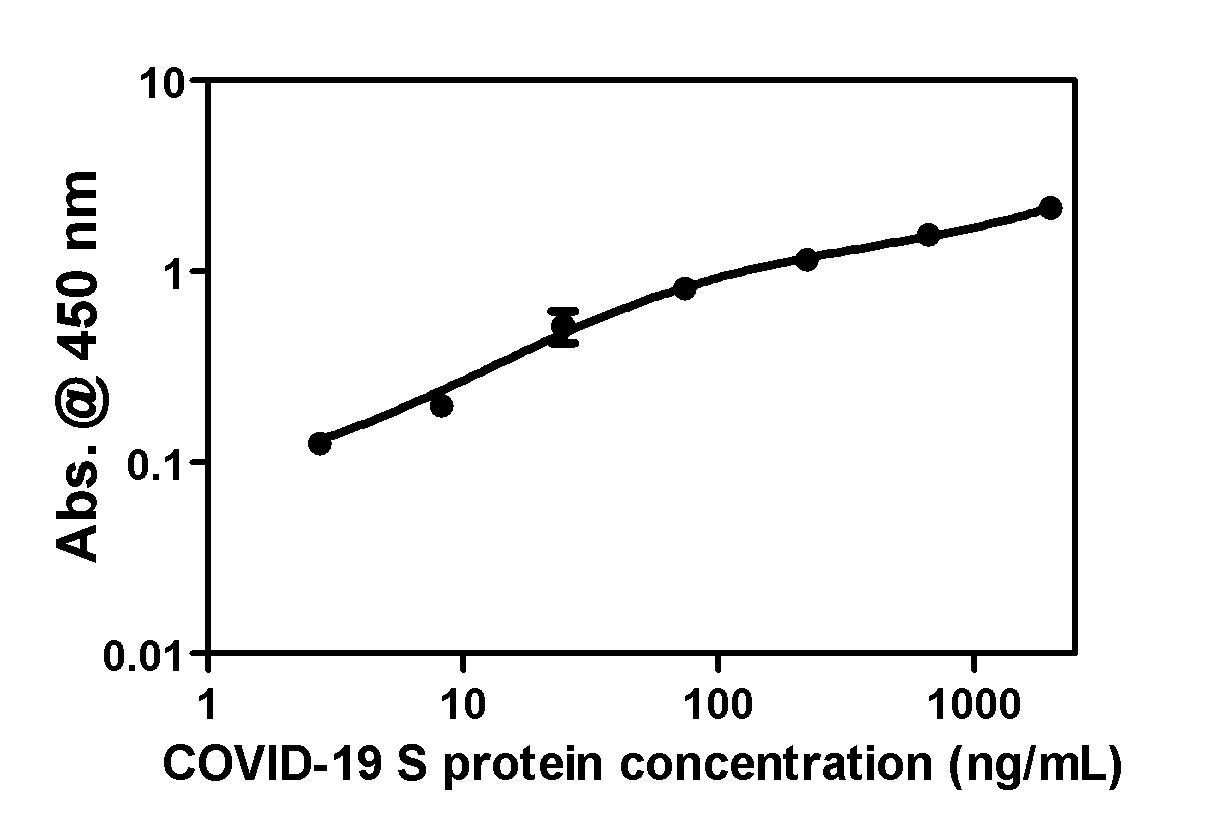

**Figure S2.** Standard curve for the determination of concentration of COVID-19 S protein.

**Experimental Procedure.**

**Detection of spike protein (S-protein) using commercial ELISA kit.**

We analyzed the collected condensate samples with a COVID-19 Spike Protein ELISA kit purchased from RayBiotech. This kit determines the presence and estimate of Spike protein (S2 subunit) of SARS-CoV-2 in the samples. and were used as per manufacturer’s protocol. Briefly, the ELISA technique was performed using a 96-well plate. Seven known concentrations (2000, 666.7, 222.2, 74.07, 24.69, 8.231, 2.744 ng/ml) of S-protein and 31 water samples) with unknown S-protein concentration were pipetted into the microliter plate wells with a volume of 100 µL of each sample. The plates were covered and incubated at room temperature (18-25 °C) for 2.5 h. After 2.5 h the wells were emptied, washed with diluted wash buffer, followed by addition of dilute biotinylated antibody (100 µL) and incubated for 1 h at room temperature. Wells were washed properly to eliminate the possibility of erroneous results. After 1h, wells were washed again with diluted wash buffer, 100 µL streptavidin added to them and incubated for another 45 min at room temperature. After 45 mins the streptavidin solution was discarded, wells were washed properly, and 100 µL of the given substrate solution mixture was added. The well plate was covered and incubated for 30 mins at room temperature in the dark with gentle shaking. After 30 mins, stop solution was added to each well. The stop solution changes the color from blue to yellow, and the intensity (absorbance) of the color was measured at 450 nm using a BioTeK plate reader and Gen 5.0 software. Measurements were tested in duplicate sets, and the average value was then utilized to determine the final S-protein concentration.

**Extraction of RNA and determination** **for the presence of SARS-CoV-2 (COVID-19) using a commercial One Step RT-qPCR kit.**

For RNA extraction, 50 mL of each heat inactivated water sample was aliquoted into sterile tubes, snap frozen and freeze dried (Freeze One 2.5, Labconco). Samples were then resuspended in 500 µL of sterile RNase free water, briefly vortexed and centrifuged at room temperature at 4000 xg for 5 minutes. They were then lysed with an equal volume of lysis buffer containing 2-mercaptoethanol and an equal volume of 100% ethanol, vortexed and added to microcentrifuge tubes fitted with spin cartridges. The samples were centrifuged at 12000 xg for 20 seconds and the flow through was discarded. They were then washed using wash buffer I and centrifuged for 20 seconds at 12000 xg and washed twice with wash buffer II containing ethanol for the same time at the same speed following the manufacture’s protocol (Invitrogen). All flow through was discarded and 200 µL of RNase free water was added to each sample and incubated for one minute at room temperature following which they were centrifuged at 12000 xg for 2 minutes and the eluent was collected. The eluent was analyzed using a NanoDrop (ThermoScientific) instrument for the presence of RNA and determination of its concentration.

If RNA was found to be present in the samples by Nanodrop measurements, then RT-PCR and RT-LAMP assay was performed to confirm the presence of COVID-19 causative virus, SARS-CoV-2. Accordingly, RT-PCR for SARS-CoV-2 (COVID-19) detection was carried out using a One Step RT-qPCR Kit from GoldBio (St. Louis, MO) run on a StepOne Plus Real Time PCR instrument (Applied BioSystems, USA). The PCR cyclic process was performed using the program detailed in the RT-qPCR protocol kit. The CDC recommended probes were used for the assay: 2019-nCoV_N1 probe, 2019-nCoV_N2 probe. TaqMan® probes are labeled at the 5′-end with the reporter molecule 6-carboxyfluorescein (FAM) and with the quencher, Black Hole Quencher 1 (BHQ-1) at the 3′-end. Briefly, 5 µL of each RNA sample was treated with 1.5 µL probe mix, 10 µL of 2X Master Mix and the volume made up to 20 µL with RNase free water. The RT-PCR was run using a first cycling step of 30 minutes at 42 °C followed by one cycle of initial denaturation with a holding time of 3 minutes at 95 °C. 40 cycles were used for denaturation and annealing/extension, the first with a holding time of 10 secs at 95 °C and the latter with a holding time of 30 seconds at 55 °C. Data was analyzed thereafter.

**One-step Loop-Mediated Isothermal Amplification (LAMP) of RNA samples.**

One-step Loop-Mediated Isothermal Amplification (LAMP) of RNA (RT-LAMP) targets was performed using WarmStart LAMP Kit (DNA & RNA) from New England BioLabs following the manufacturer’s protocol. The primers were designed by PrimerExplorer V5 software targeted for N gene segment of SARS-CoV-2. Briefly, the primer mix was prepared and 5 µL of each RNA sample was treated with 12.5 µL of the supplied 2X Master Mix, 0.5 µL of fluorescent dye (50X), 2.5 µL of the prepared primer mix (10X) and the volume made up to 25 µL with RNase free water. The samples were then incubated on a heat block with gentle shaking at 65 °C for 30 minutes followed by deactivation at 85 °C for another 5 minutes. They were then diluted 3 times and added to the wells of a microplate reader and the fluorescent emission intensity was recorded.

**Table S1.** Concentration of COVID-19 S-protein as determined by the ELISA assay.

| **Sample Number** | **Concentration (ng/mL)** |
| --- | --- |
| 0630_4 | 0.52 |
| 0630_3 | 0.56 |
| 0701_3 | Not detected |
| 0701_4 | 0.41 |
| 0702_4 | 0.55 |
| 0702_3 | 0.43 |
| 0703_3 | 0.56 |
| 0703_4 | 0.59 |
| 0630_1 | 0.26 |
| 0630_2 | 0.37 |
| 0701_2 | 0.15 |
| 0701_1 | 0.55 |
| 0702_1 | 0.42 |
| 0702_2 | Not detected |
| 0703_2 | 0.37 |
| 0703_1 | 0.63 |
| 0706_2b | 0.50 |
| 0706_4 | 0.91 |
| 0706_1d | 0.85 |
| 0706_1a | 0.94 |
| 0706_1b | 0.80 |
| 0706_1c | 1.13 |
| 0706_3 | 1.36 |
| 0706_2c | 2.61 |
| 0706_2a | 0.67 |

Note: The sample code starts with the date of sample collection from hospital followed by the dehumidifier number, i.e. 0630_4 indicates the water sample has been collected from dehumidifier number 4 on June 30, 2020.

**Table S2**. Comparison of lanthanide-doped carbon nanoparticles sensor array results with Spike protein ELISA.

| **Sample Number** | **LCNPs_biosensor (ΔI/I_0_)** | **Results based on our sensor (15 minutes)** |
| --- | --- | --- |
| 0630_4 | 0.43 | -VE |
| 0630_3 | 0.49 | -VE |
| 0701_3 | 0.46 | -VE |
| 0701_4 | 0.55 | -VE |
| 0702_4 | 0.47 | -VE |
| 0702_3 | 0.51 | -VE |
| 0703_3 | 0.52 | -VE |
| 0703_4 | 0.48 | -VE |
| 0630_1 | 0.30 | -VE |
| 0630_2 | 0.39 | -VE |
| 0701_2 | 0.26 | -VE |
| 0701_1 | 0.29 | -VE |
| 0702_1 | 0.33 | -VE |
| 0702_2 | 0.31 | -VE |
| 0703_2 | 0.47 | -VE |
| 0703_1 | 0.47 | -VE |
| 0706_2b | 0.76 | -VE |
| 0706_4 | 1.40 | -VE |
| 0706_1d | 1.66 | -VE |
| 0706_1a | 2.38 | +VE |
| 0706_1b | 2.97 | +VE |
| 0706_1c | 2.65 | +VE |
| 0706_3 | 2.18 | +VE |
| 0706_2c | 2.37 | +VE |
| 0706_2a | 1.95 | -VE |

Note: The sample code starts with the date of sample collection from the hospital followed by the dehumidifier number, i.e. 0630_4 indicates the water sample has been collected from dehumidifier number 4 on June 30, 2020.

**Table S3.** Summarization of RNA extraction results for the sampling period between July 22–August 10, 2020.

| **Sample Number** | **Concentration (ng/µL)** | **A_260_/A_280_** | **A_260_/A_230_** | **Inference** |
| --- | --- | --- | --- | --- |
| 0723_1a | - | - | - | No RNA detected |
| 0723_2a | - | - | - | No RNA detected |
| 0723_3a | - | - | - | No RNA detected |
| 0723_4a | - | - | - | No RNA detected |
| 0728_1a | 0.4 | 1.23 | 0.09 | RNA detected |
| 0728_2a | 0.3 | 1.8 | 0.07 | RNA detected |
| 0728_3a | 0.4 | 1.39 | 0.19 | RNA detected |
| 0728_4a | 0.5 | 1.50 | 0.05 | RNA detected |
| 0730_1a | - | - | - | No RNA detected |
| 0730_2a | - | - | - | No RNA detected |
| 0730_3a | - | - | - | No RNA detected |
| 0730_4a | - | - | - | No RNA detected |
| 0803_1a | - | - | - | No RNA detected |
| 0803_2a | 0.1 | 0.57 | 0.01 | RNA detected |
| 0803_3 | 2.7 | 1.36 | 0.25 | RNA detected |
| 0803_4 | 0.3 | 0.33 | 0.01 | RNA detected |
| 0807_1a | 1.3 | 1.72 | 0.01 | RNA detected |
| 0807_2a | 0.3 | 0.57 | 0.06 | RNA detected |
| 0807_3a | 0.6 | 0.79 | 0.02 | RNA detected |
| 0807_4 | 0.2 | 0.37 | 0.00 | RNA detected |
| 0810_1a | 0.2 | 0.24 | 0.08 | RNA detected |
| 0810_2a | 0.2 | 0.32 | 0.03 | RNA detected |
| 0810_3a | - | - | - | No RNA detected |
| 0810_4 | - | - | - | No RNA detected |

Note: The sample code starts with the date of sample collection from hospital followed by the dehumidifier number, i.e. 0723_1a indicates the water sample has been collected from dehumidifier number 1 on July 23, 2020.

**Table S4.** Results of RNA extraction from samples collected in VTM.

| **Sample Collection Date, Number** | **Concentration (ng/µL)** | **A_260_/A_280_** | **A_260_/A_230_** | **Inference** |
| --- | --- | --- | --- | --- |
| 0903_1a | - | - | - | No RNA detected |
| 0903_2a | 6.2 | 1.52 | 0.03 | RNA detected |
| 0903_3a | - | - | - | No RNA detected |
| 0903_4a | 0.1 | 0.24 | 0.0 | RNA detected |
| 0910_1a | 2.0 | 1.48 | 0.14 | RNA detected |
| 0910_2a | 12.5 | 1.51 | 0.58 | RNA detected |
| 0910_3a | 1.5 | 1.15 | 0.05 | RNA detected |
| 0910_4a | 0.9 | 0.67 | 0.15 | RNA detected |

Note: The sample code starts with the date of sample collection from hospital followed by the dehumidifier number, i.e. 0903_1a indicates the water sample has been collected from dehumidifier number 1 on September 3, 2020.
